# Developmental deviation in delay discounting as a transdiagnostic indicator of risk for child psychopathology

**DOI:** 10.1101/2022.10.04.22280697

**Authors:** Jacob DeRosa, Keri Rosch, Stewart H. Mostofsky, Aki Nikolaidis

## Abstract

**Background:** The tendency to prefer smaller, immediate rewards over larger, delayed rewards is known as Delay Discounting (DD). Developmental deviations in DD may play a key role in characterizing psychiatric and neurodevelopmental disorders. Recent work provided empirical support for DD as a transdiagnostic process in various psychiatric disorders. However, there is a lack of research relating developmental changes in DD from mid-childhood to adolescence to psychiatric and neurodevelopmental disorders.

**Methods:** The current study addresses this gap in a robust psychiatric sample of 1843 children and adolescents aged 5-18 (M = 10.6, SD = 3.17; 1219 males, 624 females). General Additive Models (GAMs) characterized the shape of age-related changes in monetary and food reward discounting for nine psychiatric disorders compared to neurotypical youth (NT; n=123). We found that over 40% of our sample possessed a minimum of at least three psychiatric or neurodevelopmental disorders. We used bootstrap-enhanced Louvain community detection to map the underlying comorbidity patterns impacting DD. We derived five subtypes based on diagnostic categories present in our sample. DD patterns were then compared across each of the subtypes. Further, we evaluated the effect of cognitive ability, emotional and behavioral problems and total household income (THI) in relation to DD across development.

**Results:** Higher discounting was found in six out of the nine disorders we examined relative to NT. DD was consistently elevated across development for most disorders with the exception of depressive disorders, with age-specific DD differences compared to NTs. Community detection analyses revealed that one comorbidity subtype consisting primarily of Attention-Deficit/Hyperactivity Disorder (ADHD) Combined Presentation and anxiety disorders displayed the highest overall emotional/behavioral problems and greater DD for the food reward. An additional comorbidity subtype composed mainly of ADHD Predominantly Inattentive Presentation, learning and developmental disorders showed the greatest DD for both food and monetary rewards compared to the other subtypes. Notably, this subtype had general deficits in reasoning ability, given their low performance on cognitive and academic achievement measures. Additionally, for this ADHD-I and developmental disorders subtype, THI was related to DD across the age span such that participants with high THI showed no differences in DD compared to NTs, while participants with low THI showed significantly worse DD trajectories than all others. Our results also support prior work showing that DD follows non-linear developmental patterns.

**Conclusions:** We demonstrate preliminary evidence for DD as a transdiagnostic marker of psychiatric and neurodevelopmental disorders in children and adolescents. Comorbidity subtypes provide insight into heterogeneity in DD across disorders and offer a unique way to identify high-risk individuals. Importantly, our findings suggest that DD is strongly related to overall intellectual reasoning ability and that, among those with lower intellectual reasoning, DD is particularly heightened in children from households with lower THI. The findings suggest that self-regulation may be particularly impaired in individuals with psychiatric and neurodevelopmental disorders with low household income.

## 1. Introduction

### 1.1 Overview

Delay discounting (DD) is a phenomenon in which individuals prefer smaller, immediate rewards over larger, delayed rewards (Green & Myerson, 2010). Recent work has identified DD as a key transdiagnostic marker across a range of psychiatric and neurodevelopmental disorders, including Attention-Deficit/Hyperactivity Disorder (ADHD), anxiety, and Autism Spectrum Disorder (ASD) (Amlung et al., 2019), such that individuals with these disorders tend to show higher discounting rates compared to neurotypical (NT) controls (Pinto et al., 2014; Sohn et al., 2014). Elucidating the specific shared behaviors and cognitive mechanisms underlying the transdiagnostic nature of DD has been a priority for psychiatric research. These differences can help inform current transdiagnostic treatment efforts (Pasion & Barbosa, 2019) for targeting core DD behavioral processes and provide markers for assessing response to intervention.

### 1.2. DD as a Transdiagnostic Marker

Higher discounting in psychiatric and neurodevelopmental disorders is well-documented (Lempert et al., 2019) and is proposed as a potential endophenotype for many problematic behaviors. DD also may be an efficient marker of individual differences relevant to treatment outcomes (Ahn et al., 2011). Yet, mixed findings have been reported on DD as a reliable transdiagnostic process (Bailey et al., 2021). A detailed understanding of the developmental variance of DD (Anandakumar et al., 2018) and how it relates to psychiatric and neurodevelopmental disorders across development is key to understanding the utility of DD as a transdiagnostic marker in developing psychiatric populations. Proper characterization of such developmental patterns would require evaluating a large and enriched transdiagnostic sample.

### 1.3. Comorbidity and Heterogeneity

Heterogeneity in cognitive profiles and behavioral presentations within disorders has been recognized as an intricate problem hindering progress in psychiatric and cognitive research (Allsopp et al., 2019). Directions to address this issue have called for hybrid approaches to identify subtypes capable of better explaining outcomes and advancing treatment-based tools (Feczko et al., 2019). Approximately 50% of mental health problems are established by age 14 and 75% by age 24, and the lifetime prevalence of two or more disorders was found to be between 17-27.7% (Kessler et al., 2005). ADHD co-occurs with depression in about 20-30% of patients, anxiety in over 25% of patients, and learning disorders in approximately 45% of patients (Larson et al., 2011; Michielsen et al., 2013). Individuals with co-occurring disorders such as ADHD, anxiety, and depression have been shown to have higher symptom severity and lower overall quality of life (Yang et al., 2013). Despite the prevalence of comorbidities, most studies have not assessed DD in psychiatric comorbidity, limiting our understanding of how underlying comorbidities contribute to DD. It is crucial to address if children and adolescents with certain comorbid disorders are at a higher risk for DD to aid our understanding of the proper development of treatment for comorbid youth.

### 1.4 Socio-economic indicators of Delay Discounting and Psychiatric illness

Household income, an indicator of social-economic status (SES), has been a robust predictor for individual differences in DD (Hampton et al., 2018). Lower household income has been correlated with a higher prevalence of mental health problems and disorders, such as depressive disorders and substance use (Zimmerman & Katon, 2005). Previous work has also shown that economic poverty has been associated with higher DD in psychiatric disorders, above and beyond SES (Lorant et al., 2007). Individuals experiencing significant financial duress are more likely to devalue delayed rewards given the need for immediate access to rewards, such as money (Oshri et al., 2019).

### 1.5 Current Study

DD was assessed using five different monetary items and one food item that were combined with factor analysis to extract factors of DD behavior (Koffarnus & Bickel, 2014). These factors were analyzed with a highly enriched sample of children and adolescents from the Healthy Brain Network Biobank (Alexander et al., 2017) to investigate two core aims.

#### Aim 1

Our first aim assessed transdiagnostic age-specific deviations of the most common psychiatric and neurodevelopmental disorders compared to those of neurotypicals (NTs) in multiple types of DD through developmental pattern modeling with Generalized Additive Models (GAMs). No previous studies have used these models to investigate within-group comparisons of DD as a function of chronological age.

#### Aim 2

Our second aim generated transdiagnostic subtypes using community detection to determine first how certain disorders cluster together and then whether DD differed across transdiagnostic subtypes and development. Finally, we investigated if household income is related to DD across these subtypes.

## 2. Methods

### Sample

#### 2.1 Healthy Brain Network (HBN) Sample

Data were obtained from the ongoing Child Mind Institute HBN Biobank (release 9.0), leveraging a community-based self-referral recruitment model to construct a transdiagnostic sample of 10,000 participants. The sample included in the following analyses consists of 1843 (430 female) participants between the ages of 5-18 (mean 10.16 ± 3.17). Participants included in the present work were based upon complete data available for the ADT-5.

### Measures

#### 2.2 Delay Discounting

The ADT-5 obtained individual discount rates by measuring k for six unique items. This task directly measures the k value for a given item. Higher k values indicate greater DD (Odum, 2011). The ADT-5 presents a series of questions between some amount of a delayed item and half that amount available immediately. These amounts remain stagnant while the delay to the larger amount is adjusted to determine the k value. The first-choice trial is always between the amount of the item delayed 3 weeks and the amount of the same item available immediately. Depending on the choice made by the participant, the delay either adjusts up (delayed choice) or down (immediate choice) by 8 delays (index 8 or 24) for the next choice trial. This continues for five-choice trials, with the delay index adjusting by an amount half that of the previous adjustment. This results in 32 potential k values (2^5^) nearly evenly spaced (on a logarithmic scale) between 1 hour and 25 years, the same number of possible indifference points at each delay of the adjusting amount procedure above. Participants completed this task six times for different commodities and delayed amounts. These included three versions where the delayed amount was altered: $10 delayed (vs. $5 now), $1,000 delayed (vs. $500 now), and $1,000,000 delayed (vs. $500,000 now). Fourth, participants completed a version presenting choices between $1,000 delivered at some point in the past and $500 delivered 1 hour ago. Fifth, an “explicit zero” version was also included, which presented choices between $1,000 delayed and $500 now, but with the options presented differently. In this version, delayed options were presented as “$1,000 in [delay] and $0 now” vs. “$500 now and $0 in [delay].” Sixth, a version of the task was completed presenting choices between 10 servings of the participant’s preferred snack food delivered after a delay vs. 5 servings now. Additional information on the ADT-5 can be found in the supporting information (S3.1).

#### 2.3 Psychiatric diagnosis

Participants and their parents or legal guardian met with a licensed clinician who administered the Kiddie Schedule for Affective Disorders and Schizophrenia (K-SADS; (J. Kaufman et al., 1997); a semi-structured DSM-V-based psychiatric interview to derive a clinical diagnosis (if applicable; (S1.1).

#### 2.4 Academic Achievement and Intelligence Measures

Participants were administered the Wechsler Individual Achievement Test, 3^rd^ edition (WIAT-III; (D. Wechsler, 2005), the Wechsler Intelligence Scale for Children, 5^th^ edition (WISC-V; for participants ages 6-17 years old; (David Wechsler, 2012) and *The Kaufman Brief Intelligence Test, Second Edition (KBIT-2*; for participants ages 5-6 years old; (A. S. Kaufman & Kaufman, n.d.). The age-adjusted standardized index and subtest scores were examined as indicators of cognitive and academic abilities (*WISC: Visual Spatial, Verbal Comprehension Fluid Reasoning, Working Memory, Processing Speed; WIAT: Numeracy, Pseudoword decoding, Spelling, Oral Expression, Listening Comprehension, Word Reading, Reading Comprehension, Math Problem Solving*). (S3.3-5).

#### 2.5 Parent and Self-Report questionnaires

Questionnaires relating to behavior, financial status, and demographics were completed by participants and their parents or legal guardians over the course of their visits. Subtest T-scores (adjusted for age and sex) from the Child Behavior Checklist (CBCL; (Achenbach, 2001) were examined as indicators of emotional and behavioral problems (EBP).

### Analysis

#### 2.6 Factor Analysis

Factor analysis was used to investigate latent choice DD constructs on the six commodities’ log-transformed k-values through an Exploratory Factor Analysis (EFA). Confirmatory Factor Analysis (CFA) was used to verify our factor structure using the R package “lavaan” (Rosseel, 2012) (S2.5-S.2.6).

#### 2.7 Community Detection

Bagging enhanced Louvain community detection (LCD; Blondel et al., 2008; Nikolaidis et al., 2022, 2021) was used to obtain data-driven diagnostic subtypes to better address the high comorbidity found within our DXs of interest. Our previous work has demonstrated the ability of bagging to enhance the signal-to-noise ratio of clustering and improve within and between sample reproducibility of clusters (Nikolaidis et al., 2020) (S2.4). All remaining DXs that were not within our primary DXs were coded under a created binary category (1 for any additional diagnoses, 0 for no additional diagnoses), dubbed “other”, which included communication, elimination, and motor developmental disorders.

#### 2.8 Developmental Analysis with Generalized Additive Models

GAMs were used to characterize age-related effects, along with diagnostic group and other demographic measures on the three DD factors using the “mgcv” package (Wood, 2018) in R. The first derivatives of the smooth function of age from the GAM model were calculated using finite differences to test for windows of significant change between groups across age. They then generated a simultaneous 95% confidence interval of the derivative (Simpson, 2018). Intervals of significant differences were identified as areas where the simultaneous confidence interval of the derivative does not include zero. Study site and sex were included as covariates in all GAMs. Follow-up GAMs covaried for Total household income (THI) and IQ, and missing data were handled via model-wise deletion.

We first compared NTs to individual diagnostic (DX) groups of interest and based on their proportion relative to the rest of the sample, including the following: ADHD Combined Presentation (ADHD-C), ADHD Inattentive Presentation (ADHD-I), anxiety disorders, depressive disorders, learning disorders, Obsessive-Compulsive Disorder (OCD), Oppositional Defiant Disorder (ODD), and ASD. GAMs with ADHD-C and ADHD-I did not include individuals with ASD. Individuals who had a comorbid diagnosis of ASD and ADHD-C or ADHD-I were assigned to their own groups, respectively. We then computed a series of GAMs that investigated our DX transdiagnostic subtypes compared to NTs.

THI was divided into bins (high and low to moderate) to investigate how economic status shapes DD patterns across transdiagnostic subtypes. The first GAMs compared DX subtypes within the high THI brackets. The second GAMs compared DX subtypes within the low/moderate THI groups. Our final analyses compared THI groups within individual subtypes to assess the economic impact. In these models, subtypes were individually extracted from the sample, and the group comparisons were between the THI high and low/moderate groups.

## 3. Results

### 3.1 Preliminary Analysis

No outliers were detected for the DD items, and missing data (n=210) were excluded before analyses. Distributions of the k-values obtained from the ADT-5 assessment were negatively skewed and given a natural log transformation. ADT-5 performance did not significantly differ between the three data collection sites. Correlations among the six DD items (range = .18 - .49) indicated that discounting at different magnitudes generally overlapped but at varying effect sizes (Figure S1).

### 3.2 Factor Analysis

Parallel Analysis “scree” plot estimation (Horn, 1965) from the R package “paran”, informed the three-factor solution (Figure S2), and EFA (Table S1) investigated the underlying factor structure of the six ADT-5 items. CFA was evaluated with the comparative fit index (CFI 0.974), the Tucker Lewis index (TLI = 0.952), and the root-mean-square error of approximation (RMSEA = 0.06) and Omega (ω = 0.84); all indicating the three-factor model to be an appropriate fit. Factor one (smaller sooner monetary reward; SSMR) revealed positive loadings from the k-values of $10, $1,000, $1000 Explicit 0, and $1,000 Past (range = .56 to .73). Factor two (Snack) loaded selectively on the snack item and Factor 3 on the $1,000,00 item (larger later monetary reward; LLMR). This three-factor solution was consistent with previous research that used dimensionality reduction as an exploratory method to detect underlying decision-making constructs (Koffarnus & Bickel, 2014; Anouk Scheres et al., 2010). The remaining analyses presented in this paper use the three derived DD factors. Correlations between the DD factors and Age, IQ, THI, SES can be seen in Figure S1.

#### Aim 1

##### 3.3 Individual Diagnostic Group GAMs

Our first set of GAMs controlled for sex and study site effects resulting in significant age-constant differences for the ADHD-C group compared to NTs on the SSMR (*t*=2.839, *p*=.005, age-window of significant differences [AWD]=7.44 - 17.88) (Figure 1A). For the snack factor, depressive disorders by age interaction emerged (*F*=3.389, *p*=.033, AWD=9.57-14.16) (Figure 1D) whereas constant effects across age were observed for youth with learning disorders (*t*=2.146, *p*=.032, AWD=5.04-17.88) (Figure 1C), anxiety disorders (*t*=1.973, *p*=.049, AWD=5.1-17.81; Figure 1B), and ADHD-I (*t*=1.995, *p*=.046, AWD=5.1-17.88) compared to NTs. Age-constant significant differences were observed for the LLMR factor among the ADHD-C (*t*=2.301, *p*=.022, AWD=5.06-17.88) (Figure 1E) and ASD without ADHD groups (*t*=2.035, *p*=.043, AWD=5.1-17.81; Figure 1F), such that both diagnostic groups showed higher DD compared to NTs. Statistical comparisons for all individual DX group GAM models can be found in Tables S2-S5.

**Figure 1.**
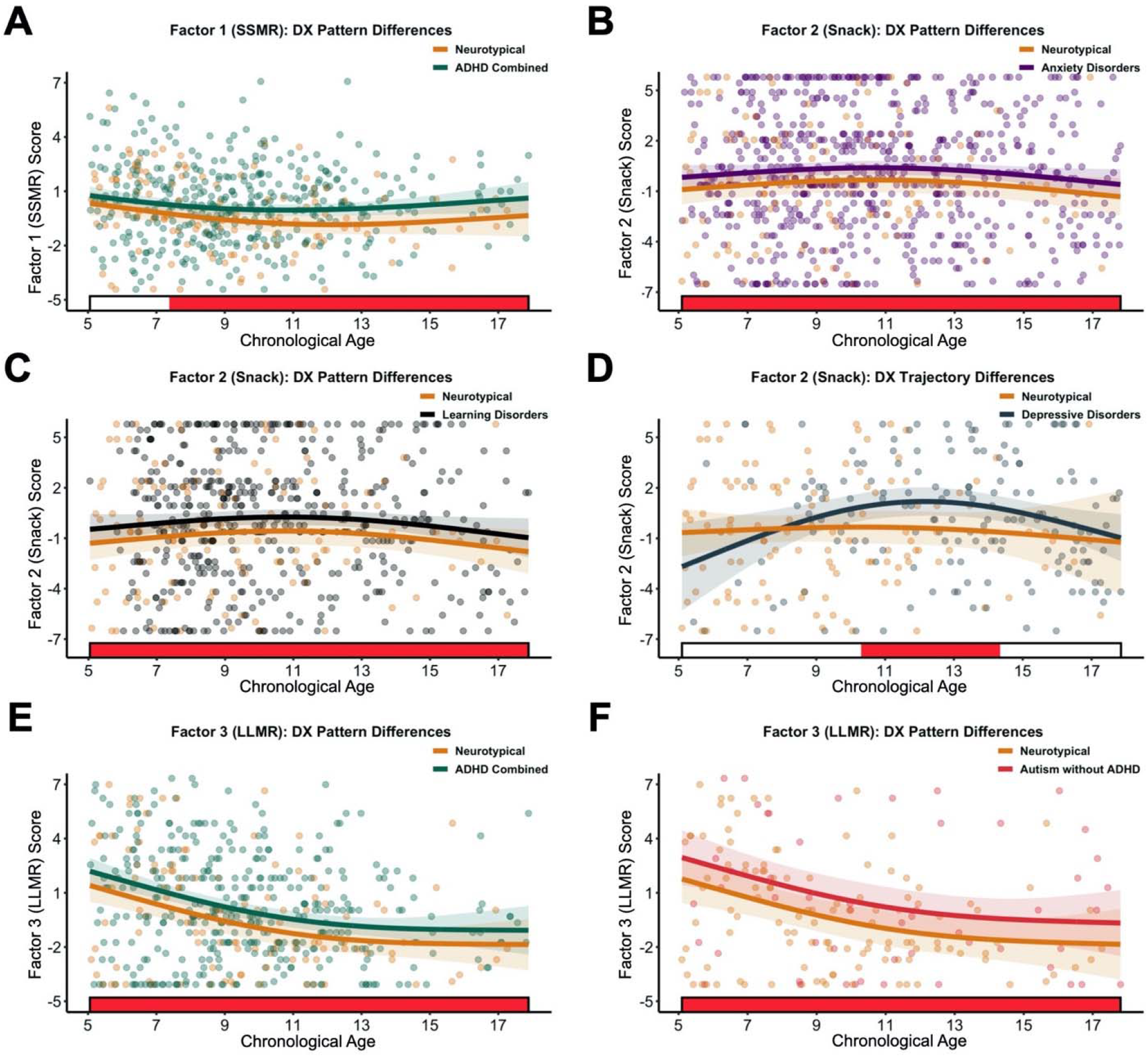
DX vs. Neurotypical DD Patterns. Red shaded bars indicate significant differences across chronological age, where the simultaneous confidence interval of the DD derivative does not include zero between groups. Lincs arc colored by group and represent the fitted GAM DD trajectory across chronological age. Points are colored by group and represent a given individual’s original DD factor scores. A) Neurotypical (orange) compared to ADHD Combined (green) on Factor 1 - SSMR. B) Ncurotypicals compared to Anxiety disorders (purple) on Factor 2 - Snack. C) Ncurotypicals compared to Learning disorders (black) on Factor 2 - Snack. D) Ncurotypicals compared to Depressive disorders (navy blue) on Factor 2 - Snack. E) Neurotypicals compared to ADHD Combined (green) on Factor 3 - LLMR. F) Ncurotypicals compared to Autism without ADHD (red) on Factor 3 - LLMR.

#### Aim 2

##### 3.4 Transdiagnostic Subtype Profiles

Bagging enhanced LCD on the binary DX categories revealed five distinct DX transdiagnostic subtypes (Figure 2). Subtype names were determined based on their primary diagnostic composition and remarkable cognitive/academic (abbreviated “Cog”) and emotional/behavioral (abbreviated internalizing [Int], externalizing [Ext], and high overall emotional/behavior problems [EBP]) profiles.

**Figure 2.**
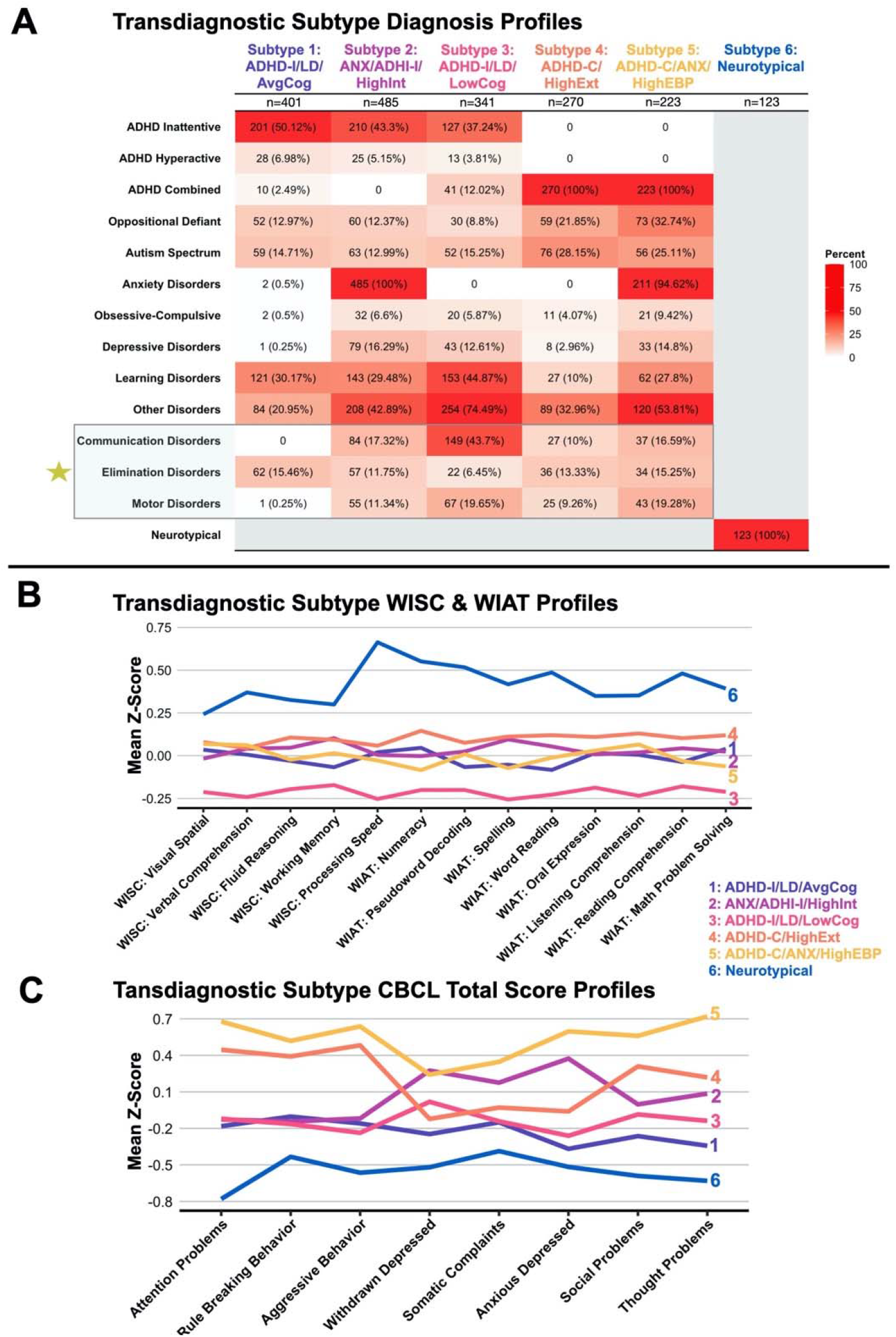
Transdiagnostic Subtype Profiles. A) LCD-derived transdiagnostic clinical diagnosis compositions Numbers represent the total count and percentage of individuals with that given diagnosis w ithin each subtype. Cells arc colored by percentage (darker red indicates higher diagnosis rates within the subtype). Note: The summed total percentage of the diagnosis cells in each subtype column will exceed 100 due to individuals with multiple diagnoses For example, every individual in subtype 2 has an Anxiety disorders diagnosis which resulted m the percentage of Anxiety disorders within that subtype equal 100. B) Mean WISC and WIAT subscale standardized scores by subtype C) Mean CBCL Total subscale standardized scores by subtype In both B and C. lines arc colored by Subtype association (ADHD-l/LD^AvgCog. light purple, ANX’ADHD-1’ Highlnt, purple; ADHD-l.’LD’lxiwCog, pink; ADI ID-C.HighExt. orange; ADHD-C.’ANX/HighEBP, yellow; Ncurotypicals. blue). Note: Gold star indicates that Communication. Elimination, and Motor disorders were not included in the original LCD analysis but were selected for inclusion in this graph for display purposes.

*Subtype 1: ADHD-I/LD/AvgCog*, was proportionally high in ADHD-I and learning disorders, with average cognitive/academic scores and relatively low to moderate emotional/behavioral problems.

*Subtype 2: ANX/ADHD-I/HighInt* was proportionally high in ADHD-I and anxiety disorders, with average cognitive/academic scores and high internalizing problems.

*Subtype 3: ADHD-I/LD/LowCog* was proportionally high in “other” disorders, learning disorders, and ADHD-I with low cognitive/academic scores and low to moderate emotional/behavioral problems.

*Subtype 4: ADHD-C/HighExt* was proportionally high in ADHD-C, with generally average cognitive/academic scores and high externalizing behavior, thought, and social problems.

*Subtype 5: ADHD-C/ANX/HighEBP* was proportionally high in ADHD-C and anxiety disorders, with average cognitive/academic scores and high emotional/behavioral problems.

One-way ANOVA tests revealed significant differences between subtypes on the CBCL, WISC, and WIAT subscales (Tables S6-S7). On average, we observe that NTs revealed the highest scores across the WISC and WIAT subscales and the lowest scores across CBCL subscales. Significant differences in sex (χ^2^ = 92.05, *p* < .001), Age (χ^2^ = 67, *p* < .001), THI χ^2^ = (22.04, *p* < .001), and Race (χ^2^ = 35.16, *p* < .05) were observed across transdiagnostic subtypes (Table 1).

**Table 1.**
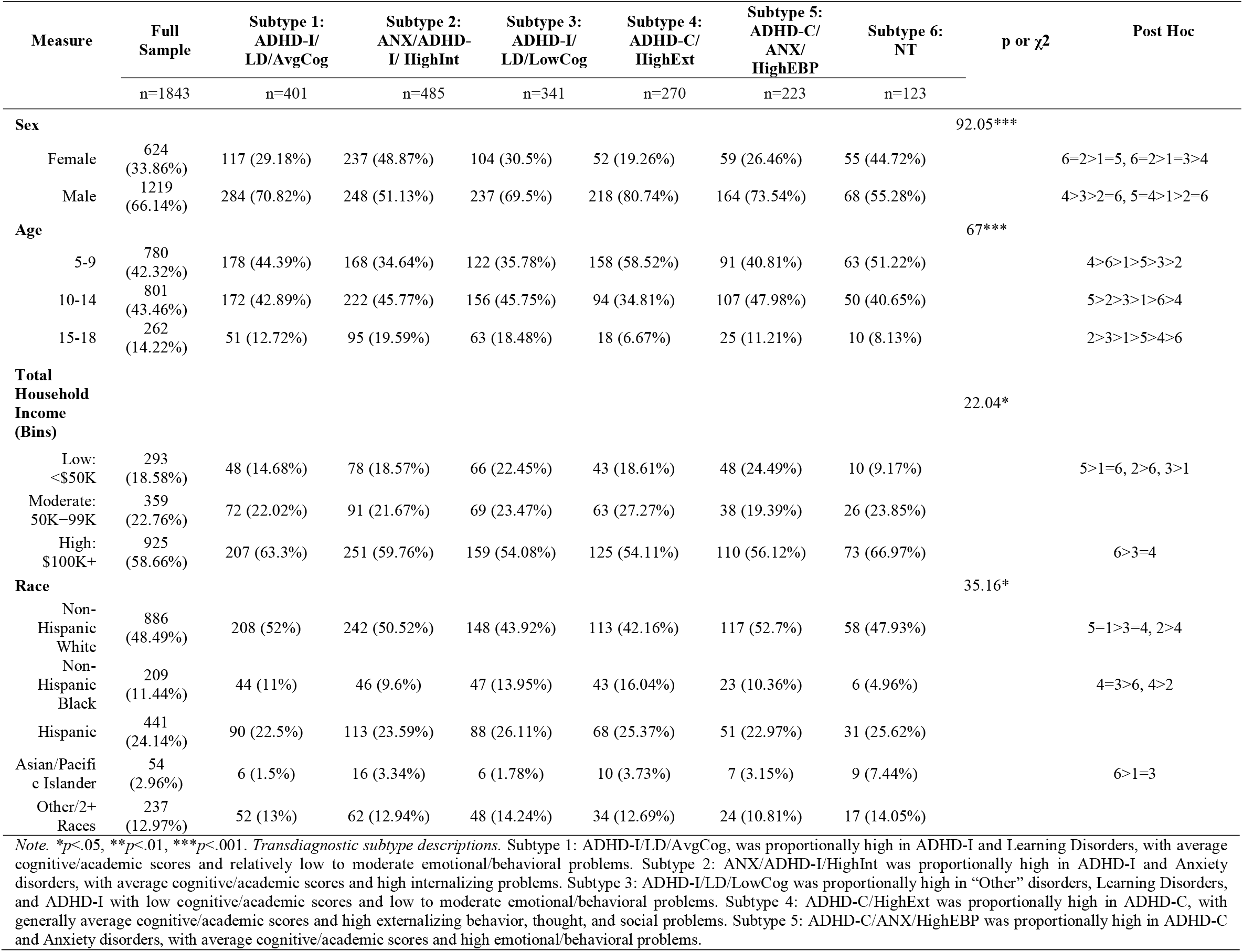
Full Sample and Transdiagnostic Subtypes Demographics.

Post-hoc z-tests indicated that relative to NTs, ADHD-I/LD/LowCog and ADHD-C/ANX/HighEBP were proportionally higher in the <50k Total Household income bracket. NTs were also proportionally higher in the $100k+ bracket compared to ADHD-I/LD/LowCog and ADHD-C/HighExt and were proportionally lower in Non-Hispanic Black populations compared to those subtypes. Relative to the other transdiagnostic subtypes, ANX/ADHD-I/HighInt was characterized by a significantly higher proportion of Females, whereas the ADHD-C/HighExt and ADHD-C/ANX/HighEBP subtypes were characterized by a significantly higher proportion of males. See Table 1 for all significant demographic post-hoc comparisons.

#### 3.6 Subtyping GAMs

GAMs tested if diagnostic subtypes show differential DD compared to NTs across development (Tables S8-S11). ADHD-I/LD/LowCog showed significantly higher DD for the SSMR (t=-1.99, p=.47, AWD=8.68-13.59; Figure 3A) and the Snack (t=-2.33, p=.02, AWD=5.1-17.88; Figure 3B), across development, whereas a group-by-age interaction was revealed for the LLMR (F=2.876, p=.017, AWD=10.74-13.75; Figure 3D). ADHD-C/ANX/HighEBP showed higher discounting across development than NTs on the snack (t=-2.529, p=.012, AWD=5.1-17.59) (Figure 3C), which remained present across all covariate models.

**Figure 3.**
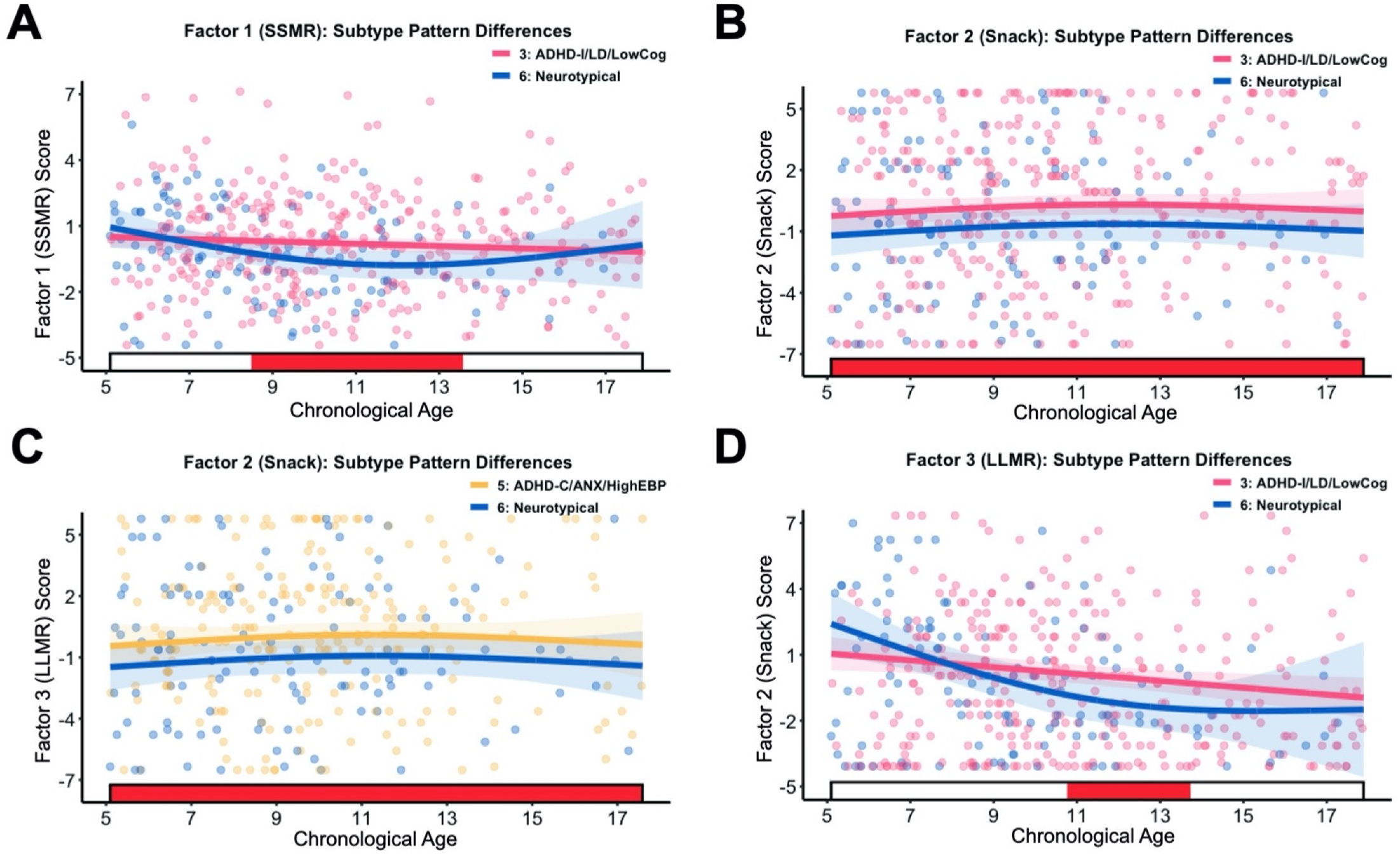
Transdiagnostic Subtype GAMs. Red shaded bars indicate significant differences across chronological age. where the simultaneous confidence interval of the DD derivative does not include zero between groups. Lines are colored by group and represent the fined GAM DD trajectory across chronological age. Points are colored by group and represent a given individual’s original DD factor scores. A) Subtype 3: ADHD-I/LD/LowCog (pink) compared to Subtype 6: Neurotypicals (blue) on Factor 1 - SSMR. B) Subtype 3: ADHGD-b’l.D.’lxiwGog (pink) compared to Subtype 6: Ncurotypicals (blue) on factor 2 - Snack. C) DX Subtype 5: ADHD-C/ANX/HighEBP (yellow) compared to Subtype 6: Ncurotypicals (blue) on Factor 1 − Snack. D) Subtype 3: ADHD-I/LD/LowCog (pink) compared to Subtype 6: Neurotypicals (blue) on Factor 3 - LLMR.

#### 3.7 Total Household Income GAMs

THI measures the total income obtained each year by both primary caregivers or one primary caregiver if only one is present in the household. Notably, THI was a significant covariate in all the monetary factor models that compared the DD pattern of ADHD-I/LD/LowCog to the NTs and the other transdiagnostic subtypes. Post-hoc analyses examined how THI differences impacted DD across subtypes.

The first pair of GAMs compared ADHD-I/LD/LowCog to the rest of the sample for all individuals with low-moderate THI. The low-moderate THI sample comparisons (SSMR: t=3.003, p=.003, AWD=5.1-17.88; LLMR: t=3.095, p=.002, AWD=8.59-17.88) (Figure 4C-D) revealed differences between ADHD-I/LD/LowCog and the rest of the sample. However, no significant differences were observed in the high THI sample subtype comparisons (Figure 4A-B). Based on the significant differences observed across our THI analyses above, we probed to evaluate if THI impacted individuals within the ADHD-I/LD/LowCog subtype. These analyses revealed that the low-moderate THI group had significantly higher DD across development compared to the high THI group (SSMR: *t*=-3.992, *p*<.001, AWD=5.22-17.88; LLMR: *t*=-4.27, *p*<.001, AWD=6.93-17.88).

**Figure 4.**
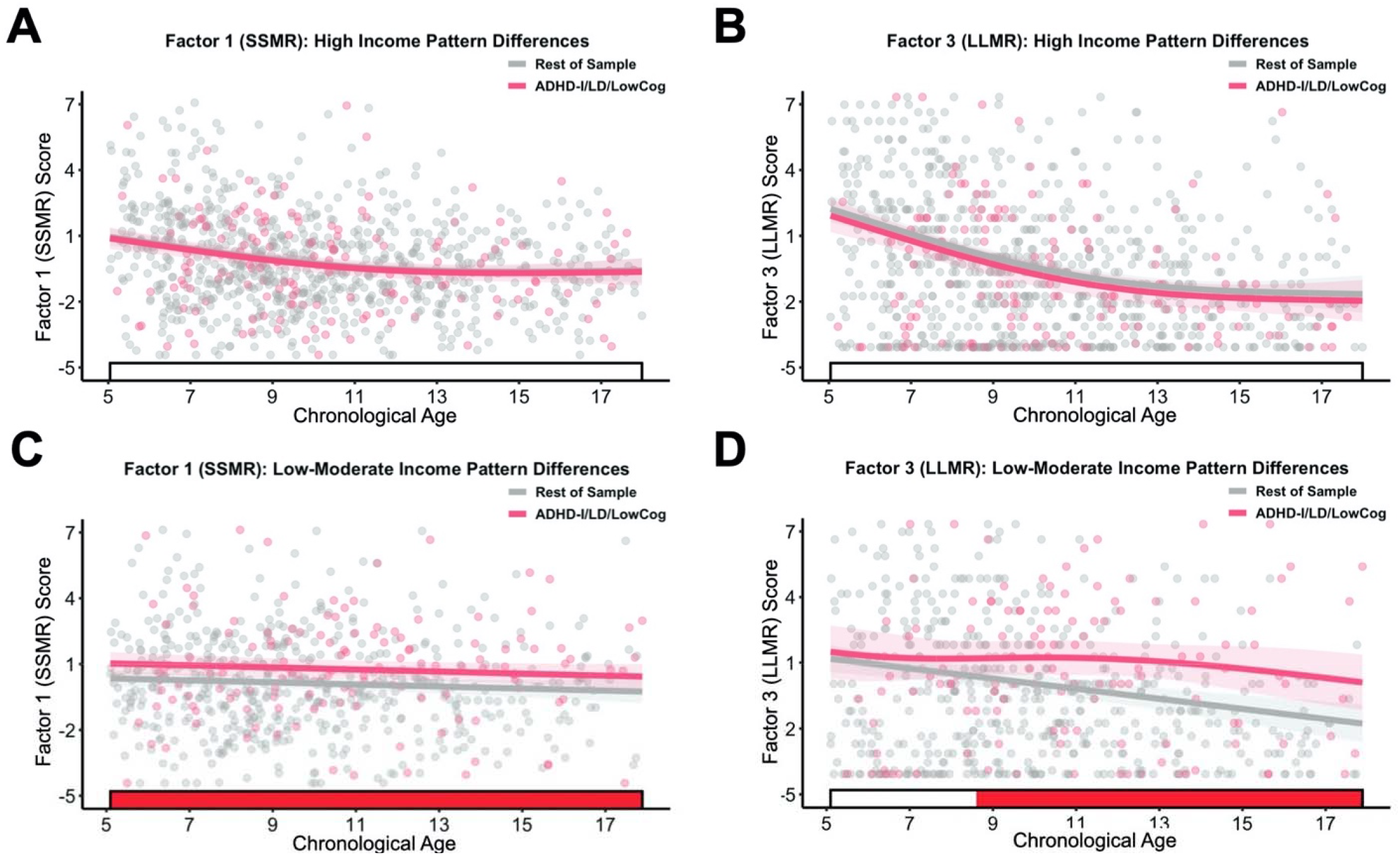
Transdiagnostic Subtype Total Household Income GAMs. Red shaded bars indicate significant differences across chronological age, where the simultaneous confidence interval of the DD derivative docs not include zero between groups. Lines are colored by group and represent the fitted GAM DD trajectory across chronological age. Points arc colored by group and represent a given individual’s original DD factor scores. Individuals were extracted from the sample based on if they had a household income higher than 100K (High) or if they were in a total household income below 99K (Low-Moderate). A) ADHD 11.D LowCog in the “High” income group (pink) compared to the rest of the sample in the “High” income group (grey) on Factor I-SSMR. B) DX ADHD-l/LD/Ix>wCog in the “High” income group (pink) compared to the rest of the sample in the “High” income group (grey) on Factor 3 - LLMR. C) ADHD-I/LD/LowCog (pink) in the “Low-.Moderate” income group compared to the rest of the sample in the “Low-Moderate” income group (grey) on Factor 1-SSMR. D) C) ADHD-I/LD/LowCog (pink) in the “Low-Moderate” income group compared to the rest of the sample in the “Low-Moderate” income group (grey) on Factor 3 - LLMR.

## 4. Discussion

### 4.1. Key Results

We first assessed the degree to which DD is a developmentally sensitive transdiagnostic process among children between the ages of 5-18 years. Compared to NTs, individuals with ADHD-C, ADHD-I, ASD without ADHD, depressive, anxiety, and learning disorders displayed higher DD for monetary or snack DD rewards. Our transdiagnostic subtypes offered the ability to address underlying heterogeneity across our diagnostic sample, revealing problematic transdiagnostic DD patterns in two distinct subtypes that included individuals with ADHD relative to NTs. Additionally, we found that household income is a critical risk factor to consider when examining monetary-based decision-making. Further, we find that lower THI exacerbates DD patterns in individuals with lower cognitive abilities. Overall, the modeling framework we present here allowed for a more precise characterization of DD as a transdiagnostic process across psychiatric and neurodevelopmental disorders and the important role of socioeconomic factors.

#### Aim 1

##### 4.2. Differences in DD across Individual Diagnostic Categories and Development

Our findings of elevated DD for monetary rewards in children with ADHD-C compared to NTs are consistent with prior work (Jackson & MacKillop, 2016; Marx et al., 2021; Patros et al., 2017) and extend this work to examine developmental changes and the impact of comorbidity. For individual diagnostic group comparisons, children with ADHD showed elevated DD for monetary rewards across development, suggesting that this may be a relatively stable trait within this population. In contrast, elevated DD was not observed in ADHD-I relative to NT for monetary rewards. This is consistent with limited prior work reporting that DD was specifically related to hyperactive/impulsive symptoms rather than inattentive symptoms (A. Scheres et al., 2008). However, children with ADHD-I did show elevated DD for food rewards that persisted across development. Given the lack of studies examining DD for non-monetary rewards across development, it will be important to replicate these findings. However, it may suggest that reward processing abnormalities are present across both the ADHD-C and ADHD-I presentations, but they may be commodity specific. Elucidating the task parameters that may lead to distinct patterns of DD in ADHD subgroups may be important for differential intervention approaches.

Elevated DD among individuals with anxiety disorders was specific to the food rewards. One issue relating this finding to previous work is that few studies exist on anxiety disorders and DD in developmental populations. One study investigating a sample of 44 adults with Social Anxiety Disorder (SAD) did not find elevated monetary DD compared to NTs (Steinglass et al., 2017). Examining the major types of anxiety disorders, such as SAD, was out of the scope of the present work. However, given the lack of research on DD within the major types of anxiety disorders, it is worth investigating how these types may differ in their developmental DD patterns across the different rewards.

Mixed findings have been reported regarding atypical DD in relation to depressive disorders (Pulcu et al., 2014) and there is still a lack of research investigating developmental changes in DD within this population. Here we find that individuals with depressive disorders show increased DD for the food reward only specific to the developmental period beginning around late childhood and early adolescence. This difference compared to NTs peaked at 11.5 years old and began to level off after 14. Importantly, depressive disorders were the only disorder that showed non-constant differences in DD across development. This suggests that for individuals with depressive disorders, the transition from childhood to adolescence may be hallmarked by periods of elevated reward processing abnormalities.

Our finding of elevated DD for monetary rewards among youth with ASD without comorbidity of ADHD is consistent with prior work that found individuals with only ASD discounted rewards more steeply than NTs and comorbid ASD with ADHD groups (Chantiluke et al., 2014). The elevated DD we observe for individuals with ASD may be due to their inability to effectively evaluate the magnitude of the LLMR due to certain cognitive deficiencies. The LLMR requires abstract prospective reasoning to consider the reward amount of $1,000,000 at delays of up to 25 years in the future. This notion aligns with previous findings on time-based and prospective processing deficiencies in individuals with ASD (Szelag et al., 2004; Williams et al., 2013). There is also a lack of research that has examined DD and learning disorders across development. Since learning disorders share a high prevalence of comorbidity with ADHD, individuals with these comorbidities may be driving the elevated DD. This notion seems to be confirmed by the elevated DD in one of our transdiagnostic subtypes, which consisted predominantly of ADHD-I and LD.

We did not observe differences in DD between individuals with OCD and NTs. Though mixed findings have been reported with OCD and DD, most studies show that individuals with OCD do not show elevated DD compared to NTs (Norman et al., 2017; Steinglass et al., 2017; Vloet et al., 2010). Finally, no differences in DD were discovered for individuals with ODD compared NTs. There is a lack of literature pertaining to ODD and DD across development; however, previous work did not find elevated DD in children with ODD and comorbid diagnosis and ADHD (Antonini et al., 2015).

Most diagnoses showed stable DD differences across development compared to NTs, except for depressive disorders whose DD patterns showed higher variability across development. Overall, our findings across these disorders support DD as a developmentally stable transdiagnostic process spanning internalizing and externalizing disorders. However, since these disorders are highly comorbid, alternative subtyping approaches are needed to clarify the most relevant clinical features/presentations across these disorders.

#### Aim 2

##### 4.3. Transdiagnostic Subtypes

DD findings among our transdiagnostic subtypes have important implications for understanding transdiagnostic processes across these disorders. DD and self-regulation may not be specific to a particular disorder but rather a subset of individuals with particular comorbidities and more severe psychopathology. Nested heterogeneity across our disorders indicates the importance of considering the effect multiple diagnoses can have on influencing individual differences in cognitive and behavioral functioning within a given diagnostic category.

Our findings reveal two distinct transdiagnostic subtypes that display elevated DD relative to NTs. The subtypes with ADHD-I, learning disorders, and low cognitive/academic scores and with ADHD-C, anxiety disorders, and high emotional/behavioral problems both displayed elevated DD for food and monetary rewards (marginal effect for the latter group) compared to the NT group and did not significantly differ from each other in DD. Together, these subtypes provided key insights into how certain individuals will display elevated DD based on their multiple comorbidity profiles. Specifically, among youth with ADHD, co-occurring affective and learning/cognitive problems may contribute to DD. It is worth mentioning that individuals with ASD may have contributed to the lack of significant differences we observed in DD with other transdiagnostic subtypes. Further work will be needed to understand the influence of ASD comorbidities in relation to their interaction with DD patterns across development.

Overall, our findings raise important questions as to whether different neurobehavioral processes contribute to elevated DD in these distinct subtypes. For example, perhaps deficits in cognitive reasoning abilities contribute to DD in the ADHD-I/LD/LowCog group, whereas increased sensitivity to immediate reward or a heightened aversion to delay and related negative affect contributes to DD in the ADHD-C/ANX/HighEBP group. Similarly, distinct neural mechanisms may contribute to elevated DD in these transdiagnostic subgroups. These findings lay the foundation for future studies with neuroimaging data to elucidate the relevant neurobehavioral processes at play. Furthermore, clinicians and researchers should consider these findings when addressing severity and heterogeneity within ADHD populations.

##### 4.4. Associations with Sociodemographic Factors

We also found that transdiagnostic subtypes were significantly associated with sociodemographic, cognitive/academic, and behavioral indices that are important to understand in the context of the above findings. First, all transdiagnostic groups had lower cognitive and academic test scores (WISC and WIAT) compared to the neurotypical group. This is consistent with increasing evidence of cognitive deficiencies in major psychiatric disorders (Goodkind et al., 2015). Intellectual reasoning ability (IQ) was also negatively associated with DD, consistent with prior research (Shamosh & Gray, 2008), providing further support for the role of intellectual reasoning abilities in reward-based decision making. In addition, we captured a subtype (i.e., ADHD-I/LD/LowCog) with higher cognitive/academic impairments with no remarkable emotional/behavioral problems compared to the rest of the sample. Their DD patterns were consistent with prior work linking cognitive abilities and self-regulation (Hofmann et al., 2012). Examining the impact of THI within this subtype notably revealed critical insights into how socio-economic hardship might have a more detrimental effect on individuals with lower cognitive abilities.

Second, we found transdiagnostic group differences in racial composition and THI. In particular, the ADHD-I/LD/LowCog and ADHD-C/HighExt subtypes contained higher proportions of individuals from minority backgrounds and were also characterized by higher proportions of individuals from the lower THI households. This finding aligns with similar developmental large-scale studies investigating these populations (Lichenstein et al., 2022) and prior evidence of a positive association between household income and mental disorders and an increased risk of maladaptive outcomes (Ballenger, 2012) that may be the result of health disparities rooted in mental healthcare (McGuire & Miranda, 2008).

The higher DD we observed in lower THI households was specific to the monetary factors and to the ADHD-I/LD/LowCog subtype, supporting previous work that implicated economic constraint with lower executive functioning and self-regulation (Oshri et al., 2019). This indicates that self-regulation toward monetary-based decision-making begins at an early age, and children living under financial constraints tend to place greater value on smaller, immediate rewards and devalue larger, delayed rewards. It may be that this is ultimately adaptive for their financial situation, considering that delayed rewards are less certain, so perhaps opting for the sooner, guaranteed reward is adaptive in a low THI context. Given the associations with this type of behavior and multiple maladaptive outcomes (MacKillop et al., 2011), it is paramount for intervention-based strategies (Amagir et al., 2018) to target these at risk-populations.

Third, there was evidence of differential representation of males and females among the DX transdiagnostic subtypes. Specifically, a higher proportion of females was present in the ANX/ADHD-I/HighInt subtype consistent with prior work linking sex differences in anxiety disorders and inattention to be more common in females than impulsivity (Quinn & Madhoo, 2014). In contrast, the ADHD-C/HighExt subtype was characterized by proportionally higher males, consistent with previous work that found boys with ADHD had higher levels of hyperactivity symptoms and externalizing behaviors (Hasson & Fine, 2012).

Finally, we found that age was negatively associated with DD, supporting established studies showing that DD is age-sensitive (Bixter & Rogers, 2019) and is sensitive to developmental changes in self-regulation and impulsivity (Orgeta, 2009). Importantly, we generally did not see evidence of differential changes in DD across development as a function of psychopathology, with the exception of depressive disorders as discussed above.

##### 4.5. Limitations

While the current work possesses many strengths, the ADT-5 was based on hypothetical rewards, which may rely more heavily on abstract reasoning than if concrete, tangible rewards were used, particularly for younger children, and may not reflect real-world DD. However, prior work in adults has shown that hypothetical rewards produce DD behavior akin to real currency (Koffarnus & Bickel, 2014). In addition, we could not assess validity in decision-making during this task. Finally, this study is limited by our sample size’s low racial and ethnic diversity. Given the previous findings on DD differences across racial and ethnic groups (Hampton et al., 2018), future studies should examine developmental differences across these populations with similar approaches applied in the current work.

##### 4.6. Generalizability

This study leveraged unique modeling approaches and a large, highly heterogeneous transdiagnostic sample to accurately characterize developmental differences in DD behavior. Our findings suggest that DD is a multi-faceted indicator of psychiatric and neurodevelopmental illness and severity across children and adolescents. This work has important implications for behavioral economics, education, psychology, and neuroscience. Previous findings (van den Bos et al., 2014) suggest that distinct striatal pathways are related to DD, and structural/functional connectivity between the striatum and prefrontal cortex and subcortical areas play a role in self-regulation and impulsivity. Further work should investigate the neural mechanisms, such as functional brain network organization and cortico-striatal system maturation, to determine their role in reward-based decision-making.

Future studies should also investigate the effectiveness of tiered intervention strategies (Rung & Madden, 2018) to reduce DD using methods such as mindfulness (Scholten et al., 2019), financial literacy programs (Amagir et al., 2018), and behavioral training (Ashe & Wilson, 2020) in school and clinical settings. These interventions should specifically target individuals with lower cognitive abilities and from lower and moderate household income brackets. Extensive work is needed using the approaches outlined here to evaluate its reproducibility and to gain a more cohesive understanding of these findings and their clinical relevance. In sum, we recognize the complexity of reward-based decision-making and encourage future work to exploit transdiagnostic modeling to advance the field toward personalized medicine.

## Data Availability

All data produced in the present study are available upon reasonable request to the authors

## Ethical Considerations

The study was approved by the Chesapeake Institutional Review Board. Written informed consent was obtained from adult participants. For participants younger than 18, written consent was obtained from their guardians, and written assent was obtained from the participant.

## Acknowledgments

The work presented here was primarily supported by gifts to the Child Mind Institute from Phyllis Green, Randolph Cowen, and Joseph Healey, as well as NIMH awards U01MH099059 & R01MH091864. The funders for this project were not involved in any part of the experimental design, analysis, and interpretation of data or manuscript preparation and submission.

## Conflict of interest

The authors declare that they have no known competing financial interests or personal relationships that could have appeared to influence the work reported in this paper.

Key points and relevance

1. Delay discounting was identified as a developmentally sensitive transdiagnostic marker of psychiatric and neurodevelopmental illness
2. From a clinical practice perspective, we highlight that transdiagnostic comorbidity subtypes offer insight into heterogeneity in delay discounting across disorders and provide a unique way to identify high-risk individuals.
3. From an educational practice perspective, we show that delay discounting is strongly related to overall intellectual reasoning ability.
4. Among individuals with lower intellectual reasoning ability, delay discounting is particularly heightened in children from lower household incomes.
5. Self-regulation may be particularly impaired in individuals with psychiatric and neurodevelopmental disorders with low household incomes.

