## Supporting Information for "Developmental deviation in delay discounting as a transdiagnostic indicator of risk for child psychopathology"

**Supplemental Materials**

**S1. Phenotyping**

**S1.1. Psychiatric Diagnosis**

The clinical team (i.e., clinician, social worker (or junior psychologist), and psychiatrist (if a consultation was required) used the K-SADS in tandem with observations from clinical visits and questionnaires to yield a consensus diagnosis for each participant. The Anxiety disorders group included participants who received a current diagnosis of any anxiety disorder (i.e., Agoraphobia, Generalized Anxiety Disorder, Other Specified Anxiety Disorder, Panic Disorder, Selective Mutism, Separation Anxiety, Social Anxiety (Social Phobia), Specific Phobia, and Unspecified Anxiety Disorder). The Depressive disorders group included participants who received a current diagnosis classified as a depressive disorder (i.e., major depressive disorder, persistent depressive disorder (dysthymia), disruptive mood dysregulation disorder, and other specified Depressive disorders). The Learning Disorder (LD) group included participants who received a current diagnosis of any LD (i.e., Specific Learning Disorder with Impairment in Mathematics, Specific Learning Disorder with Impairment in Reading, and Specific Learning Disorder with Impairment in Written Expression).

**S2. Analysis Methods**

**S2.1. ADT-5 Factor Structure and Correlations**

Consistent with k, higher scores on these factors indicate higher DD. Age (range = -0.25 to -0.14), IQ (range = -0.17 to -0.14), THI (range = -0.09 to -0.08), and SES (-0.08), were significantly correlated with the SSMR and LLMR. The Snack was not significantly correlated with Age (Pearson r=0.03), THI (Pearson r=0.03), or SES (Pearson r=0.01).

Follow-up associations were then examined between the DD Factors and subscales of the WISC and WIAT (S Figure 2), and two sets of associations were conducted with the CBCL subscales - one set controlling for IQ and one without (S Figure 3.). Results indicated the DD factors were more strongly associated with the WISC (range = -.2 to -.01) and WIAT (range = -0.21 to -0.08) compared to the CBCL (range = -0.05 to 0.07). We also examined associations between SES and THI with the WISC, WIAT, and CBCL subscales. On average, higher SES and THI were associated with lower behavioral problems (range = -.16 - .05) and higher academic achievement (range = .22-.34), and intelligence scores (range = .18-.32) (S. Table 4.).

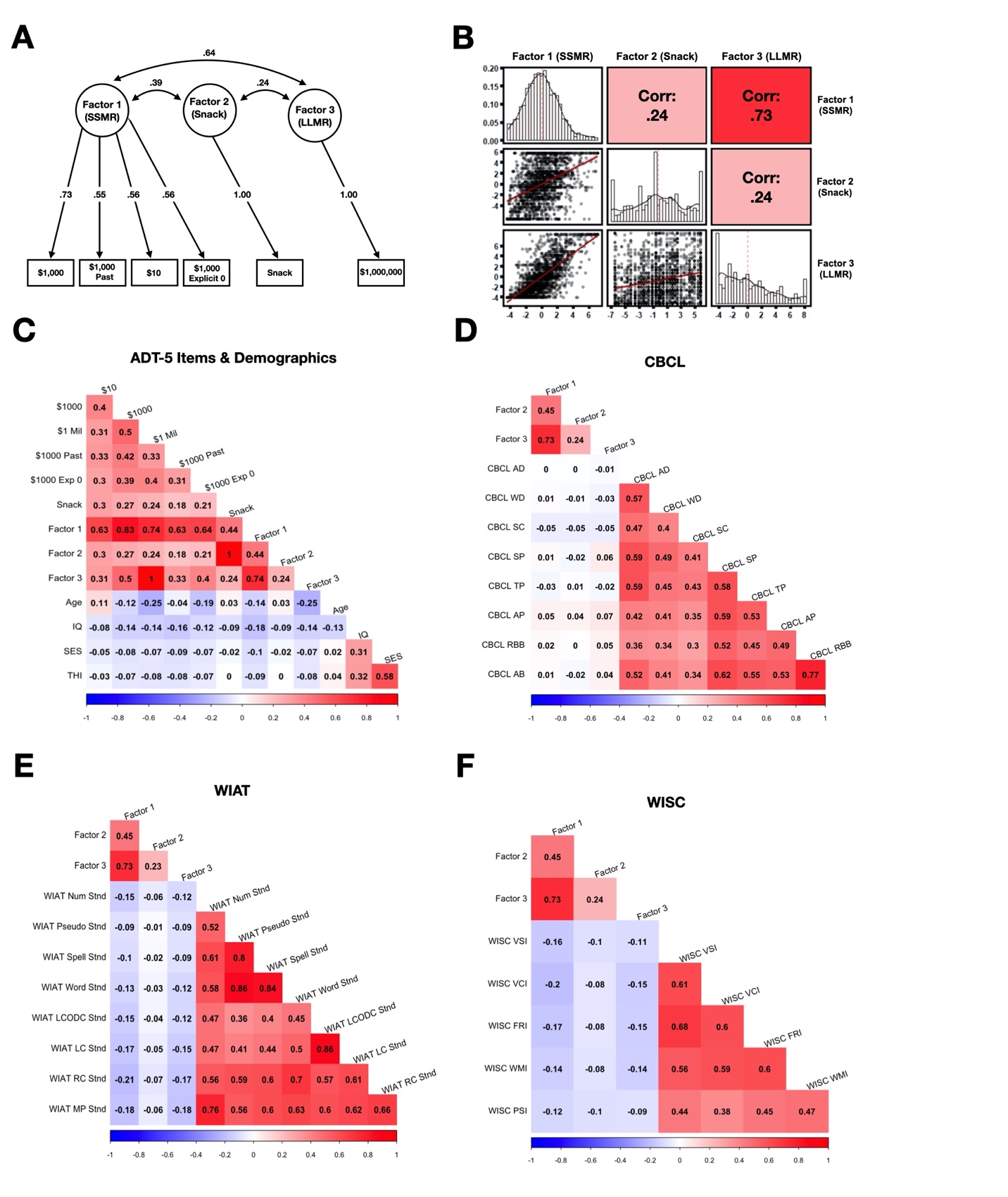

**Figure S1.** A) Factor loading structure of the individual DD items and grouped by color (Factor 1 – SSMR; Factor 2 – Snack; Factor 3 – LLMR). B) Correlation pair plot for each of the factors. The lower left plots indicate the relationship with the fitted regression line in red and individual points in black. The diagonal indicates the distribution of points across each of the factors. The upper right diagonal represents the total correlation value between each factor. C) Correlations between each of the individual DD items, DD factors, Age, IQ, SES, and Total Household Income (THI). D) Correlations between the DD factors and the eight CBCL subscale total scores (AD = Anxious Depressed, WD = Withdrawn Depressed, TP = Thought Problems, SC = somatic complaints, AP = Attention Problems, RBB = Rule-Breaking Behavior, AB = Aggressive Behavior, SP = Social Problems. E) Correlations between the DD factors and the eight WIAT subscale standardized scores. F) Correlations between the DD factors and the eight WISC subscale standardized scores.

**S2.2. Parallel Analysis**

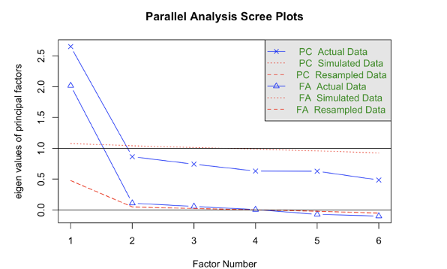

**Figure S2.** Parallel analysis “scree” plot of the successive six DD commodities’ eigenvalues. Breaks in the plot suggest the three factors as an appropriate number of components or factors to extract.

The extraction of factor analysis decomposition relies on selecting a limit to the number of dimensions to extract. Some studies suggest that whatever number of components crosses some specific threshold of percentage variance-accounted-for. Others suggest doing so through visual inspection of the scree-plot, which is the descending eigenvalue/variance-accounted-for. These approaches can lead to highly variable and non-identical results. Parallel Analysis, a non-parametric method of testing for this significance, has been developed to use a data-driven approach to selecting the number of components. Briefly, through repeated permutations of the data and subsequent calculations of the FA, one can measure the null distribution of eigenvalues of each component. Thus, providing a test for the eigenvalue of each factor.

**S2.3. Louvain Community Detection**

LCD is an iterative network analytic approach that detects sub-groups through two steps. First, the algorithm applies a "greedy" assignment of nodes to communities, favoring local optimizations of modularity. Second, the definition of a new coarse-grained network in terms of the communities found in the first step. Both steps are repeated until no further modularity-increasing reassignments of communities are possible. An attractive feature of LCD for data-driven clustering compared to other such methods (i.e., hierarchical, k-means) is the algorithmic determination of the number of clusters. This feature can help inhibit incorrect cluster approximation in cluster choice.

The algorithm inputs a matrix of N single-cell measurements and partitions them into subpopulations by clustering a graph representing their phenotypic similarity. It then builds this graph in two steps. First, it finds the k nearest neighbors for each cell resulting in N sets of k-neighborhoods. Second, it operates on these sets to build a weighted graph such that the weight between nodes scales with the number of neighbors they share. The Louvain community detection method is then used to find a graph partition that maximizes modularity. A graph's modularity (Q) is a quantitative measure of the number of edges found within communities compared against the number predicted in a random graph with an equivalent degree distribution. Positive Q values indicate that the number of intracommunity edges exceeds those predicted statistically. Q can range from −1.0 to +1.0, with 0 indicating no subgroups and 1.0 indicating a perfectly reliable division of groups. Depending on how nodes are assigned to communities, a wide range of Q may be found for a graph. The optimal community structure sought by the Louvain algorithm is the number of node assignments that returns the highest Q. An attractive feature of LCD for data-driven subtyping compared to other clustering methods (i.e., hierarchical, k-means) is its algorithmic determination of k. This feature inhibits incorrect k approximation and or researcher bias for the number of clusters to obtain.

**S2.4. Subtype Stability**

We first computed a k-nearest neighbor (KNN) graph (thus, connecting samples with a similar profile) from ten diagnostic group labels. To prevent overfitting and bolster profile robustness and reliability, K for the KNN graph was determined using the "nclass Sturges" formula, which implicitly bases k on the range of data. A weighted network was then created by applying Fisher's exact test to compute the statistical significance of the overlap in neighbors between each pair of samples. LCD was applied on the weighted network as the final step.

Bagging begins by resampling a dataset with replacement (i.e., bootstrapping) and then aggregating across bootstrap samples. This technique reduces variability in the estimation process by averaging multiple resampled datasets. When applied to clustering, bagging has been shown to improve robustness and reliability. The fundamental advantage of bagging stems from the additional value of combining multiple cluster assignments into a single clustering solution. More specifically, the features for ensemble clustering consist of the aggregation of cluster outputs themselves. LCD is then applied, resulting in cluster solutions for every bootstrap iteration. Each cluster solution is transformed into individual adjacency matrices summed together to create an adjacency matrix (similarity matrix) of the total number of times participants were in the same cluster. Next, mask adjacency matrices are simultaneously created and summed to equate the number of times participants went into the same LCD iteration together (inclusion matrix). The similarity matrix is then divided by the inclusion matrix to create a mean adjacency matrix (stability matrix) that is then turned into a weighted network. LCD is applied one more time to obtain the final cluster solution.

**S2.5. Factor Analysis**

Each unrotated factor was statistically independent of one another, which is unlikely to be the case in the data itself. To this end, a Promax rotation was applied to these factors to be correlated to one another and thereby more accurately represent the proper relationships between variables in our data. This rotation method was applied using the “Promax” function from the R package “GPArotation”.

**S2.6. Confirmatory Factor Analysis**

The DD k values between various tasks showed consistent positive associations between the different items, indicating a shared dimensional structure. K values were more similar across similarly valued reward magnitudes suggesting underlying shared variance between subject’s patterns based on the magnitude of reward (Noor, 2011). These reward sizes likely involve different decision-making mechanisms (Roesch & Bryden, 2011). Our derived 3-factor solution explained significantly more variance than would be expected due to chance, demonstrating the robustness of the underlying factor structure. CFA fit indices demonstrated high internal consistency of the factor structure, supporting the notion that the dimension reduction of the ADT-5 tasks was able to capture robust underlying factor structure. The three factors that emerged suggest that both reward size and type matter and that the responders will show dissociable patterns based on these key attributes. This was supported by the correlation structure of the data with similar reward magnitudes demonstrating higher correlations.

**S2.7. General Additive Models**

The k value was chosen for the current study because it facilitates assessing relationships between groups and differential scores. Higher k values indicate more discounting for a given DD task commodity. Previous studies have suggested that DD may be nonlinear across development Anandakumar et al., 2018). For this reason, we propose a novel approach to evaluate the intrinsic complexity of DD behavior with Generalized Additive Models (GAMs). No previous studies have used these models to investigate within-group comparisons of DD as a function of chronological age. These models augmented a multifaceted characterization of DD and its relationship with psychiatric and multimorbidity subtype outcomes.

Like a general linear model, predictors in a GAM can be replaced by smooth functions of themselves, offering an efficient and flexible estimation of nonlinear effects. Each group comparison for diagnostic category vs. NT and subtype was modeled as an ordered factor in the GAMs. In the case of ordered factors, the model fits *L-*1 differences smooths where *l=*1*,…, Ll=*1 are the levels of the factor and *L* the number of levels. These smooths model the difference between the smooth estimated for the reference level and the *l*1th level of the factor.

Individual smooths are centered for identifiability reasons. The first *s(Age)* in the model is the smooth effect of Age on the reference level of the ordered factor *Group*. The second smoother, s(Age, by = *Group*) is the set of *L*-1difference smooths, which model the smooth differences between the reference level smoother and those of the individual levels (excluding the reference). The model still estimates a separate smoother for each level of the ordered factor. The smoother for the reference level is assessed through the contribution from *s(Age)* only.

In contrast, the smoothers for the other levels are formed from the additive combination of *s(Age)* and the relevant difference smoother from the set created by *s(Age, by = Group).* This is analogous to the situation for estimating an ANOVA. The intercept becomes an estimate of the mean response for the reference level of the factor, and the remaining model coefficients estimate the differences between the mean response of the reference level and that of the other factor levels. All models in our analyses were fit with splines to capture linear or nonlinear age effects.

**S3. Sample Description**

**S3.1 ADT-5**

The task was developed to rapidly assess discount rates in a short period of time (i.e., under 5 minutes) through the presentation of five trials for each commodity and its delayed amount. A trained research assistant (RA) administered the ADT-5 to participants on a laptop during one of the study visits. Parameters for each session were automated before administration of the task. Before the ADT-5 was started, the following instructions were read to the participant:

“You will now complete a series of decision-making tasks. You will be asked to make choices between different amounts of something. There are no right or wrong answers in the tasks; just choose which option you prefer in each case. Please take your time and answer thoughtfully. To select the option on the left side of the screen, press the left arrow, and to select the option on the right side of the screen, press the right arrow.”

When the program started, a terminal window appeared and prompted the RA to enter the participant’s ID, session, and the participant’s favorite snack, and then proceeded to the first run. There was a total of six runs for the task. The first trial of each run began with the commodity delayed at three weeks and half of its value available immediately. The subsequent trials’ delay was titrated up or down dependent on the participant’s choice on the previous trial. The task provided 32 potential and k values on a logarithmic scale that ranged from one hour to 25 years for each of the six commodities.

**S3.2 Financial Status Questionnaire (FSQ)**

Internally developed, the FSQ is administered to the participant’s parent to legal guardians to assess household income, public assistance received, and health insurance information.

**S3.3 Child Behavior Checklist (CBCL)**

The CBCL is a device by which parents rate their child's problem behaviors and competencies. It consists of 118 items related to behavior problems, which are scored on a 3-point scale ranging from not true to often true of the child. The main scoring for the CBCL used confirmatory factor analysis to test the structure groupings of sets of behaviors that typically occur in tandem. Similar questions are grouped into several symptomology scale scores*,* and their scores are summed to produce a raw score for that syndrome. The eight empirically based symptomology scales were used in our analyses.

**S3.4 Wechsler Individual Achievement Test - III (WIAT)**

The WIAT is a test of academic achievement administered to all participants. It is a comprehensive yet flexible measurement tool useful for achievement skills assessment, learning disability diagnosis, special education placement, and clinical appraisal for preschool children through adults.

*Spelling.* Measures written spelling of letter sounds and single words. *Pseudoword Decoding.* Measures the ability to decode nonsense words. *Word Reading.* Measures several important areas for developing reading skills: naming letters, letter-sound correspondence (alphabetic principle), phonological awareness, and word reading comprehension. *Numeracy.* Measures untimed, written math calculation skills in the following domains: basic skills, basic operations with integers, geometry, algebra, and calculus. *Math Problem Solving*. Measures untimed math problem-solving skills in the following domains: basic concepts, everyday applications, geometry, and algebra. *Reading Comprehension*. Measures untimed reading comprehension of various types of text, including fictional stories, informational text, advertisements, and how-to passages. *Oral Expression.* The Oral Expression subtest contains three components: Expressive Vocabulary: Measures speaking vocabulary and word retrieval ability. *Oral Word Fluency:* Measures efficiency of word retrieval and flexibility of thought processes. *Listening Comprehension.* Contains two components: Receptive Vocabulary: Measures listening vocabulary. *Oral Discourse Comprehension*: Measures the ability to make inferences about and remember details from oral sentences and discourse.

**S3.5 Wechsler Intelligence Scale for Children-V**

The WISC-V is a measure of cognitive function in children and adolescents. Participants completed the ten core subtests: similarities, vocabulary, blocks, matrix, figure weights, digit span, coding, symbol search, visual puzzles, and pictures span.

*Working Memory Index:* The Working Memory Index (WMI) measured the participant's ability to register, maintain, and manipulate visual and auditory information in conscious awareness, which requires attention and concentration, as well as visual and auditory discrimination. Within the WMI, Picture Span (PS) required the participant to memorize pictures and identify them in order on subsequent pages. On Digit Span (DS), participants listened to strings of numbers read aloud and recalled them in the same order, backward order, and ascending order. The Digit Span Forward (DSf) scaled process score is derived from the total raw score for the Digit Span Forward task. The participant was required to repeat numbers verbatim on this task, with the number of digits in each sequence increasing as the task progressed. This task required working memory when the number of digits exceeded the participants ability to repeat the digits without rehearsal. This task represents essential capacity in the phonological loop.

The Digit Span Backward (DSb) scaled process score is derived from the total raw score for the Digit Span Backward task. This task invoked working memory because the participant was required to repeat the digits in a reverse sequence than was initially presented, requiring participants to manipulate the information before responding mentally. The Digit Span Sequencing (DSs) scaled process score is derived from the total raw score for the Digit Span Sequencing task. This task required the participant to sequence digits according to value, invoking quantitative knowledge in addition to working memory. The increased demands for mental manipulation of information on the Digit Span Sequencing task place additional demands on working memory and attention. In addition to the two subtests in the WMI, Letter-Number Sequencing (LN) was administered to gain a more comprehensive understanding of the participant’s working memory proficiency. On this subtest, participants were read sequences of numbers and letters, then recalled the numbers from lowest to highest and the letters in alphabetical order.

*Visual-Spatial Index.* The Visual-Spatial Index (VSI) measured the participant’s ability to evaluate visual details and understand visual-spatial relationships to construct geometric designs from a model. This skill requires visual-spatial reasoning, integration and synthesis of part-whole relationships, attentiveness to visual detail, and visual-motor integration. The VSI consists of two tasks. During Block Design (BD), the participants viewed designs and used blocks to recreate each design. Visual Puzzles (VP) required participants to view a completed puzzle and select three pieces that would reconstruct the puzzle. Regarding individual subtests within the VCI, Similarities (SI) required the participants to describe similarities between words with common characteristics, and Vocabulary (VC) required participants to name pictures and define words aloud.

*Fluid Reasoning Index.* The Fluid Reasoning Index (FRI) measured the participant’s ability to detect the underlying conceptual relationship among visual objects and use reasoning to identify and apply rules. Identification and application of conceptual relationships in the FRI require inductive and quantitative reasoning, general visual intelligence, simultaneous processing, and abstract thinking. The FRI consists of two subtests: Matrix Reasoning (MR) and Figure Weights (FW). Matrix Reasoning required the participants to select the missing piece to complete a pattern. On Figure Weights, the participant looked at a scale with a missing weight and identified the weight to keep the scale balanced.

*Processing Speed Index.* The Processing Speed Index (PSI) measured the participant’s speed and accuracy of visual identification, decision making, and decision implementation. Performance on the PSI is related to visual scanning, visual discrimination, short-term visual memory, visuomotor coordination, and concentration. The PSI assessed participant’s ability to rapidly identify, register, and implement decisions about visual stimuli. The PSI consists of two-timed subtests. Symbol Search (SS) required participants to scan a group of symbols and mark the target symbol. On Coding (CD), participants copied symbols that were paired with numbers.

*Verbal Comprehension Index.* The Verbal Comprehension Index (VCI) measured the participant’s ability to access and apply acquired word knowledge. Specifically, this score reflects the ability to verbalize meaningful concepts, think about verbal information, and expression using words.

**S3.6. Supplemental Table Descriptions**

**Table S1.** Exploratory factor analysis of the ADT-5 EFA loadings for each of the 6 DD items. Loadings with absolute values <0.40 have been removed for clarity.

**Table S2.** GAM regression table for Diagnostic Group vs NT, age, sex, and interactions. Statistics are reported as t statistics for parametric terms and F statistics for smooth terms.

**Table S3.** GAM regression table for Diagnostic Group vs NT, age, sex, IQ, and interactions. Statistics are reported as t statistics for parametric terms and F statistics for smooth terms.

**Table S4.** GAM regression table for Diagnostic Group vs NT, age, sex, THI, and interactions. Statistics are reported as t statistics for parametric terms and F statistics for smooth terms.

**Table S5.** GAM regression table for Diagnostic Group vs NT, age, sex, THI, and interactions. Statistics are reported as t statistics for parametric terms and F statistics for smooth terms.

**Table S6.** IQ, WISC, and WIAT index and subscale standardized Mean and (SD) scores by Subtype. F statistics are reported, and p values are indicated by asterisks according to their level of significance (. *p<.05, **p<.01, ***p<.001). Post-Hoc comparisons are shown based on if the subtype post-hoc z-test was significant and ordered according to what subtype was significantly greater than the other subtype(s).

**Table S7.** CBCL Total Mean and (SD) scores by Subtype. F statistics are reported, and p values are indicated by asterisks according to their level of significance (. *p<.05, **p<.01, ***p<.001). Post-Hoc comparisons are shown based on if the subtype post-hoc z-test was significant and ordered according to what subtype was significantly greater than the other subtype(s).

**Table S8.** GAM regression table for Subtype, age, sex, THI, IQ, and interactions. Statistics are reported as t statistics for parametric terms and F statistics for smooth terms.

**Table S9.** GAM regression table for Subtype, age, sex, IQ, and interactions. Statistics are reported as t statistics for parametric terms and F statistics for smooth terms.

**Table S10.** GAM regression table for Subtype, age, sex, THI, and interactions. Statistics are reported as t statistics for parametric terms and F statistics for smooth terms.

**Table S11.** GAM regression table for Subtype, age, sex, THI, IQ, and interactions. Statistics are reported as t statistics for parametric terms and F statistics for smooth terms.
